# Modeling the transmission of antibiotic-resistant Enterobacterales in the community: a systematic review

**DOI:** 10.1101/2024.01.15.24301323

**Authors:** Eve Rahbé, Philippe Glaser, Lulla Opatowski

## Abstract

**Background:** Infections by antibiotic-resistant Enterobacterales are a public health threat worldwide. While dissemination of these opportunistic pathogens has been largely studied in hospitals, less is known about their acquisition and spread in the community. Here, we aim to characterize mechanistic hypotheses and scientific contributions of mathematical modeling studies focusing on antibiotic-resistant Enterobacterales in the community.

**Methods:** We conducted a systematic review of mathematical modeling studies indexed in PubMed and focusing on the transmission of antibiotic-resistant Enterobacterales in the community (i.e., excluding models only specific to hospitals). For each study, we extracted model features (host population, setting), formalism (compartmental, individual-based), biological hypotheses (transmission, infection, antibiotic use impact, resistant strain specificities) and main findings. We discussed additional mechanisms to be considered, open scientific questions, and most pressing data needs to further improve upon existing epidemiological modeling.

**Results:** We identified 18 modeling studies focusing on the human transmission of antibiotic-resistant Enterobacterales in the community (n=11) or in both community and hospital (n=7). Models aimed at: (i) understanding mechanisms driving resistance dynamics; (ii) identifying and quantifying transmission routes; or (iii) evaluating public health interventions to reduce resistance. Studies highlighted that community transmission, compared to hospital transmission, play a significant role in the overall acquisition of antibiotic-resistant *Escherichia coli*. Predictions across models regarding the success of public health interventions to reduce resistance rates depended on pathogens, settings, and antibiotic resistance mechanisms. For *E. coli*, lowered person-to-person transmission led to greater reduction in antibiotic resistance rates compared to lowered antibiotic use in the community (n=2). For *Klebsiella pneumoniae* lowered antibiotic use in the hospital led to greater reduction compared to lowered use in the community (n=2). Finally, we reported a moderate number of modeling studies inferring parameters from empirical data (n=9), probably due to a critical lack of available data for antibiotic resistance rates in the community.

**Conclusions:** We found a limited number of modeling studies addressing the transmission of antibiotic-resistant Enterobacterales in the community, highlighting a need for model development and extensive community-based data collection. Such modeling will be critical to better understand the spread of antibiotic-resistant Enterobacterales in the community and design public health interventions specific to this setting.

## Introduction

Antibiotic resistance in clinically relevant bacteria is a major threat for global health.^1^ Yet its control, both at local and global scales, remains a challenge. Antibiotic resistance epidemiology is highly heterogeneous, as the emergence, selection and spread of resistance in human populations differs across pathogens, antibiotic resistance mechanisms and transmission settings. As a consequence, observed resistance rates vary greatly depending on the antibiotic and bacterial species under study.^2^

*Escherichia coli* and *Klebsiella pneumoniae*, two species of the order Enterobacterales, are estimated to be among the bacteria associated with the greatest burden of antibiotic resistance worldwide.^3^ Enterobacterales are frequent opportunistic pathogens, predominantly carried asymptomatically in the human gut but commonly responsible for extra-intestinal diseases, including potentially severe infections of the urinary tract, respiratory tract and bloodstream.^4^ Since the 1980s, they have been increasingly associated with resistance to successive generations of extended-spectrum *β*-lactams, later followed by carbapenem resistance.^5^ For *E. coli* alone, antibiotic resistance was associated with an estimated 829,000 deaths in 2019 (95% confidence interval: 601,000-1,120,000).^3^ For decades, antibiotic-resistant Enterobacterales infections were mostly reported as hospital-acquired, causing a large nosocomial burden.^6^ At present, significant levels of extended spectrum *β*-lactamase (ESBL)-producing *E. coli* carriage are reported in healthy populations worldwide, with highest rates observed in South-East Asia.^7,8^ In turn, a growing number of ESBL-producing *E. coli* infections are now reported as being community-acquired.^9,10^

Mathematical models of infectious disease transmission (“models”) are increasingly used in the field of antibiotic resistance epidemiology, to better characterize the emergence and transmission routes of antibiotic-resistant bacteria (ARB) in human populations, and to support public health decisions regarding ARB control.^11^ As they rely on mechanistic hypotheses, models have generally focused on the transmission of specific antibiotic-bacteria combinations within specific environments. On one hand, “hospital” models formalize ARB selection and transmission within healthcare settings. Such models have been widely used to evaluate impacts of interventions such as antibiotic stewardship and patient isolation, and to assess transmission in different medical environments such as intensive care units and long-term care facilities.^12^ The most studied ARB in hospital models are those historically at highest risk of nosocomial spread, including methicillin-resistant *Staphylococcus aureus* and to a lesser extent ESBL-producing Enterobacterales.^13^ On the other hand, “community” models address ARB spread within human populations outside healthcare settings. Most published community models have focused on antibiotic-resistant *Streptococcus pneumoniae* or *Neisseria gonorrhoeae*.^14^ While many Enterobacterales and other clinically important bacteria also circulate extensively in the community, their transmission and control in this setting has been more rarely investigated.^13,14^

The biological and epidemiological mechanisms that drive antibiotic-resistant Enterobacterales selection and transmission in the community remain relatively poorly characterized and little quantified. ^15–17^ For instance, the contribution of horizontal gene transfer (HGT) within or between different reservoirs in driving the population-level epidemiology of antibiotic resistance is still poorly understood,^18^ as is the role of asymptomatic carriage in driving transmission and infection^19,20^ (note: we use the terms carriage and colonization interchangeably). It is also unclear how bacterial persistence in the environment contributes to maintain community transmission.^15,21^ Mathematical modeling can help make sense of the scarce data available and disentangle the various mechanisms potentially contributing to observed dynamics.

Here, we systematically review published mathematical models of antibiotic-resistant Enterobacterales transmission in the community. We summarize models’ characteristics and biological hypotheses and highlight their contributions and limitations. Finally, we discuss open scientific questions and the data sources needed to support the development of new models tailored to this growing global health problem.

## Methods

### Database and search terms

We searched PubMed for published mathematical modeling studies describing the population-level human-to-human transmission of antibiotic-resistant Enterobacterales (*E. coli* and/or *K. pneumoniae*) in the community. The search included all studies presenting original results (excluding review articles) published in English between 1 January 1992 and 1 July 2023. We used the following search query:

> (“transmission model*”[Title/Abstract] OR “dynamic model*”[Title/Abstract] OR “mechanistic model*”[Title/Abstract] OR “mathematical model*”[Title/Abstract] OR “compartment model*”[Title/Abstract] OR “compartmental model*”[Title/Abstract])
>
> AND
>
> (“commensal*”[All Fields] OR “escherichia coli”[All Fields] OR “coli”[All Fields] OR “e coli”[All Fields] OR “klebsiella pneumoniae”[All Fields] OR “klebsiella”[All Fields] OR “k pneumoniae”[All Fields] OR “enterobacterales”[All Fields] OR “enterobacteriaceae”[All Fields] OR “gram-negative”[All Fields])
>
> AND
>
> (“antibiotic resistance”[Title/Abstract] OR “antimicrobial resistance”[Title/Abstract] OR “multidrug resistance”[Title/Abstract] OR “beta lactam resistance”[Title/Abstract] OR “carbapenem resistance”[Title/Abstract] OR “antibiotic-resistant”[Title/Abstract] OR “multidrug-resistant”[Title/Abstract] OR “beta lactam-resistant”[Title/Abstract] OR “carbapenem-resistant”[Title/Abstract] OR “ESBL”[Title/Abstract])

We included modeling stodies on commensal bacteria in addition to Enterobacterales, to also retrieve studies that focused on generic intestinal commensals. We did not explicitly add search terms such as “community” or “general population” in case these were too restrictive.

### Model selection: inclusion and exclusion criteria

Studies reporting a mathematical model formalizing the transmission of antibiotic-resistant Enterobacterales between human hosts in the community were eligible. We excluded studies which were in the following categories:

– *different methods*: studies not presenting a mechanistic transmission model, but rather statistical models, time-series analysis, or cost-effectiveness analysis;
– *different host population*: transmission models not focusing on humans, but rather on animals, livestock or pets;
– *different pathogens*: transmission models specifically designed for bacteria other than Enterobacterales, like *Helicobacter pylori* or *S. pneumoniae*;
– *different settings*: models only describing transmission in healthcare settings (hospitals, long-term care homes, nursing homes), without including the community;
– *different scales*: microscopic-level models (pharmaco-dynamic or pharmaco-kinetic models, microbial population models, within-host models, …) not also including populational-level transmission.

### Data extracted and qualitative analysis

In a first step, we extracted key model features including: studied antibiotic-bacteria combinations, specificities of the host population (general population, high-risk groups), data sources used for estimation of antibiotic use parameter values (country, region, theoretical), and studied transmission settings (households, community, community and hospital). We summarized model hypotheses regarding bacterial transmission, the infection process, and effects of antibiotic exposure on carriage. The potential inclusion of any additional mechanisms was also summarized, such as potential resistance-associated fitness costs, multiple resistance acquisition routes and host population structure (Table 1). Finally, models were characterized according to the type of epidemiological antibiotic resistance data used (none, aggregated or individual-level) to perform parameter estimation (Table 3).

**Table 1.** Summary of the 18 modeling studies included in the review.

| Objectives | Study | Model features |  |  |  | Model formalism | Biological hypotheses |  |  | Additional mechanisms included |  |  | Data for inference (details in Table 3) |
| --- | --- | --- | --- | --- | --- | --- | --- | --- | --- | --- | --- | --- | --- |
|  |  | Drug-bug pair * | Type of hosts | Antibiotic use data | Setting ** |  | Transmission | I vs C *** | Antibiotic impact | R strain specificities | Bacterial acquisition | Host population structure |  |
| 1. Impact of mechanisms on dynamics | Austin et al. 1997<sup>36</sup> | G | General population | Theoretical | C | Deterministic compartmental | R-S | - | R emergence | Reduced transmission | Human-to-human + background | - | - |
|  | Eisenberg et al. 2012<sup>31</sup> | APR-EC | General population | Village, Ecuador | C | Deterministic compartmental | R | - | R emergence | - | Human-to-human + background | - | - |
|  | Blanquart et al. 2018<sup>35</sup> | G | General population | Theoretical | C | Deterministic compartmental | R-S | - | Increased S clearance depending on susceptibility | Reduced transmission | Human-to-human | Yes | - |
|  | Davies et al. 2019<sup>26</sup> | APR-EC<br>FQR-EC | General population | European countries | C | Stochastic individual-based approximated by a deterministic compartmental | R-S-multi | - | Increased S clearance & R emergence | Reduced transmission | Human-to-human | - | Aggregated |
|  | Olesen et al. 2020<sup>29</sup> | G | General population | European countries | C | Deterministic compartmental | R-S-multi | - | Increased S clearance | Reduced transmission | Human-to-human | Yes | - |
|  | Godijk et al. 2022<sup>32</sup> | ESBL-EC | General population | Netherlands | C + H | Deterministic compartmental | R-S-multi | Yes | Increased S clearance & R emergence | Different resistance-associated fitness costs and benefits | Human-to-human + background | - | - |
| 2. Transmission route identification & quantification | Arcilla et al. 2017<sup>37</sup> | ESBL-E | High-risk group in household | - | C | Stochastic individual-based | R | - | No effect | - | Human-to-human + background | - | Individual |
|  | Haverkate et al. 2017 (x2 models)<sup>25</sup> | ESBL-E | High-risk group in household | - | C | Stochastic individual-based / Deterministic compartmental | R | - | No effect | - | Human-to-human + background | - / Yes | Individual |
|  | Knight et al. 2018<sup>38</sup> | ESBL-EC | General population | United Kingdom | C + H | Deterministic compartmental | R | Yes | Increased R acquisition | Reduced infection risk | Human-to-human + background | - | - |
|  | Mughini-Gras et al. 2019<sup>23</sup> | ESBL-EC | General population | - | C | Deterministic compartmental | R | - | No effect | - | Human-to-human + background | - | - |
|  | Van den Bunt et al. 2020<sup>33</sup> | ESBL-E | General population | - | C | Deterministic compartmental | R | - | No effect | - | Human-to-human | - | Individual |
|  | Allel et al. 2022<sup>39</sup> | GN | General population | Village, Chile | C + H | Deterministic compartmental | R | Yes | Increased R acquisition | - | Human-to-human + background | - | Aggregated |
| 3. Intervention evaluation | Sundqvist et al. 2010<sup>34</sup> | TrimR-EC | General population | Kronoberg, Sweden | C | Deterministic compartmental | R-S | - | Increased S clearance | Increased clearance | Human-to-human | - | Aggregated |
|  | Talaminos et al. 2016<sup>28</sup> | ESBL-EC | General population | Spain | C + H | Deterministic + stochastic compartmental | R-S | Yes | Increased R infection risk | - | Human-to-human | - | - |
|  | MacFadden et al. 2019<sup>22</sup> | ESBL-EC | General population | Sweden | C + H | Deterministic compartmental | R-S | - | Increased S clearance & R acquisition | Reduced transmission | Human-to-human | - | Aggregated |
|  | Kardas-Sloma et al. 2020<sup>30</sup> | ESBL-EC | General population | France | C | Stochastic individual-based | R | - | Increased R acquisition | - | Human-to-human + background | - | - |
|  | Kachalov et al., 2021<sup>24</sup> | 3GCR-KP<br>CR-KP | General population | European countries | C + H | Deterministic compartmental | R-S | Yes | Increased S clearance & R emergence | Reduced transmission | Human-to-human + background | - | Aggregated |
|  | Salazar-Vizcaya et al. 2022<sup>27</sup> | ESBL-KP | General population | Switzerland | C + H | Deterministic compartmental | R | Yes | Increased R acquisition | - | Human-to-human + background | - | Aggregated |

-: not included in the model
\* G: generic bacteria; GN: Gram-negative bacteria; EC: *E. coli*; KP: *K. pneumoniae*; ESBL: extended-spectrum beta-lactamase producing bacteria; TrimR: trimethoprim resistant; 3GCR: third-generation cephalosporin resistant; CR: carbapenem resistant; APR: aminopenicillins resistant; FQR: fluoroquinolone resistant \*\* C: community; H: hospital \*\*\* I vs C: infection versus carriage differentiation; S: sensitive bacteria; R: resistant bacteria

In a second step, we summarized each study’s major findings. Reviewed articles were categorized according to their main scientific goals: (i) analyzing the impact of biological and/or populational mechanisms on antibiotic resistance dynamics; (ii) identifying routes of transmission and/or quantifying transmission; and (iii) evaluating the impact of public health interventions on antibiotic resistance trends.

## Results

Our search returned 192 publications, of which 18 met inclusion criteria and were included in the review (see PRISMA flowchart in Figure 1). ^22–39^

**Figure 1.**
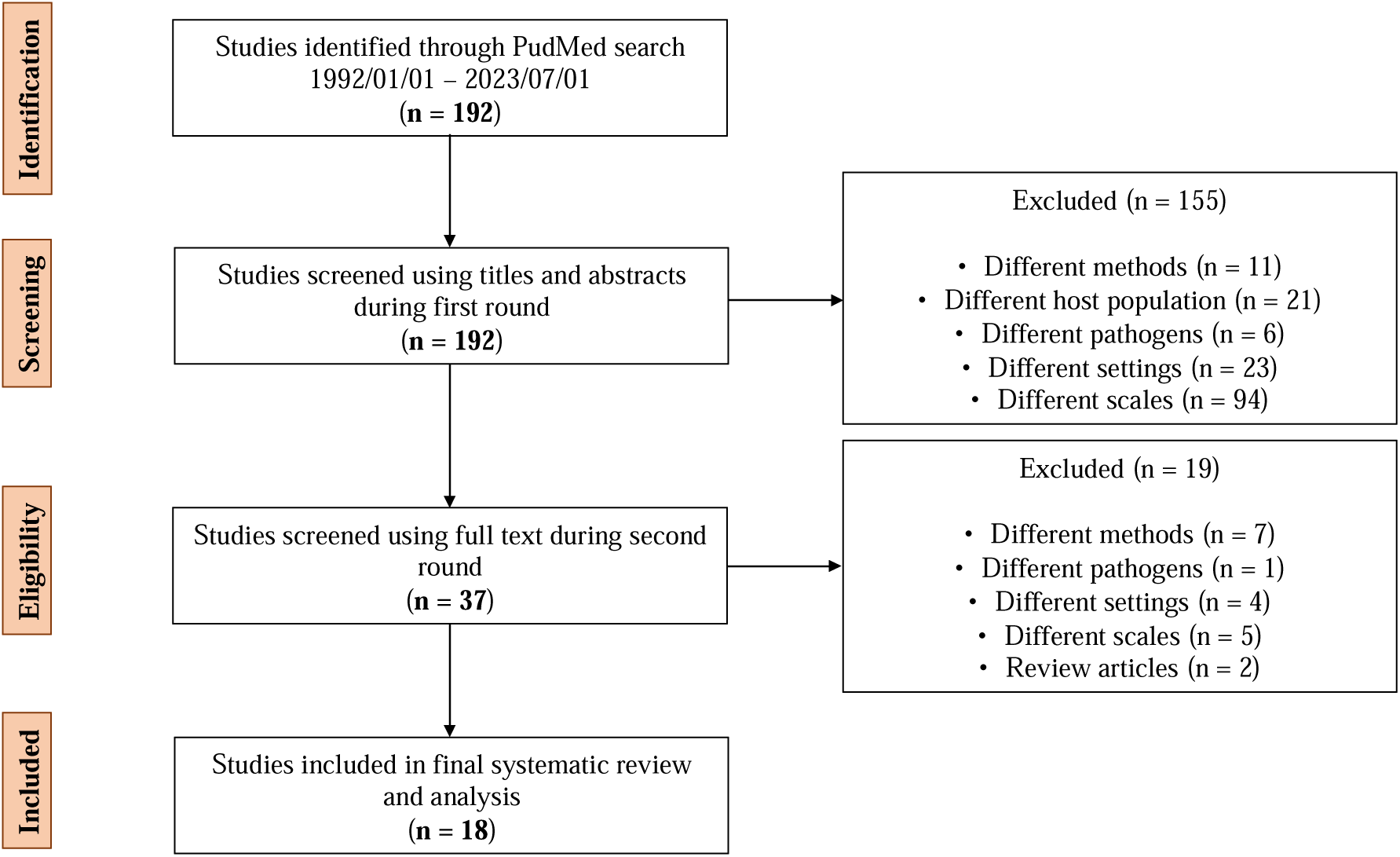
PRISMA flowchart representing screening and selection of publications included in the reviewing process.

### Model features

The included modeling studies predominantly described *E. coli* transmission (9/18, Table 1). The others modeled transmission of generic commensal bacteria (3/18), generic Enterobacterales (3/18), *K. pneumoniae* (2/18) or generic Gram-negative bacteria (1/18). ESBL production was the most frequent type of antibiotic resistance mechanism modeled (10/18), while just one model focused on carbapenem resistance, in *K. pneumoniae* (1/18).^24^ Host population was mainly representative of the general human population. Two studies specifically focused on high-risk groups, either returning travelers or returning hospitalized patients.^25,37^ Concerning antibiotic use, ten studies explicitly used parameter values obtained from high income countries (France, Sweden, Switzerland, United Kingdom, United States, or European countries), two studies focused on low-resource villages in South America (Ecuador and Chile),^31,39^ while two studies used theoretical parameters, and the remaining four did not specify antibiotic use parameters. While eleven models focused on community settings only, seven explicitly modeled both community and healthcare settings, considering setting-specific transmission parameters and antibiotic use rates. Compartmental modeling was the most frequent formalism used (n=15), while two models were individual-based, and one study included both a compartmental and individual-based model.

### Biological hypotheses

#### Between-human bacterial transmission

The between-human transmission of antibiotic-resistant Enterobacterales was modeled in three different ways in the included studies (Figure 2): (i) models formalizing the transmission of resistant strains only (“R” models); (ii) models formalizing the transmission of sensitive or resistant strains (“R-S” models); and (iii) models focusing on the transmission of sensitive or resistant strains and allowing for single or multi-carriage of strains (“R-S-multi” models). Multi-carriage models have been proposed in the field of antibiotic resistance mathematical modeling to reproduce the observed coexistence of resistant and sensitive strains across different levels of antibiotic use, both at the individual- and population-level.^40^

**Figure 2.**
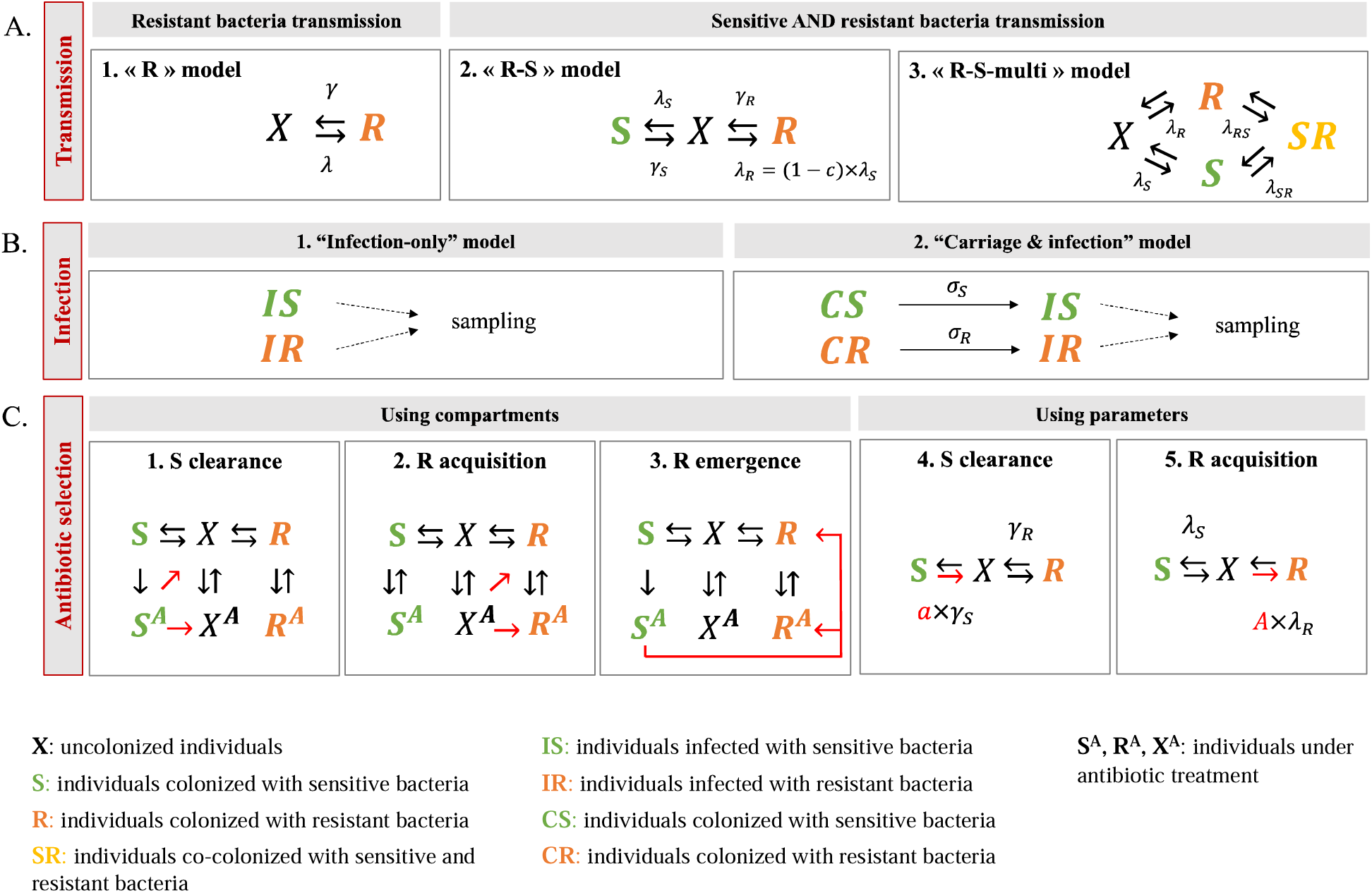
Hypotheses of mathematical models regarding between-human bacterial transmission, bacterial carriage and infection, and impacts of antibiotic use on the population-level dynamics of antibiotic resistant Enterobacterales. (A) Transmission. 3 mechanisms considered for between-human bacterial transmission: transmission of resistant strains only (1, “R”); transmission of both sensitive and resistant strains (2, “R-S”); and transmission of sensitive and resistant strains with both single and multi-carriage compartments (3, “R-S-multi”). **(B) Infection.** 2 mechanisms considered for bacterial carriage and infection: model only considering infection compartments (1, “infection-only”); and model accounting for both carriage and infection compartments, using a progression-to-disease rate between the two (2, “carriage & infection”) – in both cases, sampling is often performed on infections. **(C) Antibiotic selection for resistance.** 3 mechanisms considered for the selective impact of antibiotic use on resistance, using different models’ formalisms with either compartments (1,2,3) or parameters (1,2): *S clearance*. the clearance of sensitive bacteria compared to no clearance or slower clearance of resistant ones (1); *R acquisition*. the increased probability of resistant bacteria acquisition (2); and *R emergence*. within the host, populations of sensitive bacteria become resistant (3). The latter can be interpreted as representing the acquisition of antibiotic resistance genes from co-colonizing microbiota through horizontal gene transfer, and/or endogenous acquisition, i.e. the outgrowth of sub-dominant resistant bacteria present, prior to antibiotic exposure, in small quantities among individuals primarily carrying sensitive strains. It is important to note that the formalisms shown here are not exhaustive. **(A-B-C) Parameter description. λ**: force of 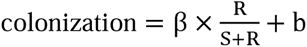; **β**: between-human transmission rate in [/ day] or [/ carriers / day]; **b**: constant background rate that captures transmission from non-human sources, horizontal transmission, or de novo emergence in [/ day]; **c**: resistance-associated fitness cost; **1/γ**: duration of carriage (1/**γ**_S_ vs 1/**γ**_R_ if sensitive and resistant bacteria have different durations) in [day]); **σ_S_,σ_R_**: progression-to-disease rate in [/ carriers / day]; **a**: multiplicative factor to increase clearance of sensitive bacteria, when individuals are under antibiotic use (a > 1); **A**: multiplicative factor to increase transmission of resistant bacteria, when individuals are under antibiotic use (A > 1).

#### Bacterial infection

While six models differentiated between asymptomatic carriage and symptomatic infection (“carriage & infection” model, Figure 2), most models (14/18, Table 1) did not (“infection-only” model, Figure 2). Although carriage is a precursor for many infections,^41,42^ its exclusion from most modelling studies likely stems from a lack of robust epidemiological data describing asymptomatic carriage of antibiotic-resistant Enterobacterales in healthy populations.

#### Antibiotic selection for resistance

Most models assumed that antibiotic exposure exerts selection pressure advantaging antibiotic-resistant bacteria (n=14). This was modelled according to three distinct mechanisms: (i) *S clearance*. the effective clearance of sensitive bacteria compared with no or less efficient clearance of resistant ones, giving the latter a relative transmission advantage; (ii) *R acquisition*. an increased probability of resistant bacteria acquisition in antibiotic exposed individuals, for instance due to loss of colonization resistance following microbiome dysbiosis; and (iii) *R emergence*, an increased probability of within-host populations of sensitive bacteria becoming resistant subsequent to antibiotic exposure, through antibiotics favoring the horizontal transfer of resistance genes from co-colonizing microbiota, or again through antibiotics killing sensitive microbiota, favoring the survival and outgrowth of resistant bacteria initially present within the host in undetectably small quantities, a phenomenon known as endogenous resistance acquisition (Figure 2C). Antibiotic use was formalized either explicitly, i.e. considering distinct compartments for treated and untreated individuals, or through multiplicative coefficients increasing rates at which sensitive strains are cleared or resistant strains are acquired.

### Main findings

The 18 reviewed modeling studies were classified according to their main scientific objectives (Table 1).

#### Analyzing the impact of biological and/or populational mechanisms on antibiotic resistance dynamics

Six modeling studies aimed to analyze how biological or populational mechanisms, such as within-host dynamics and host population structure, impact antibiotic-resistant Enterobacterales transmission dynamics (Table 1). Five out of these six studies used the “R-S” or “R-S-multi” model structures.

Several modeling studies formalized within-host mechanisms to investigate their impact on host-level antibiotic resistance dynamics. Austin et al. first explored how resistance-associated fitness costs in bacterial transmission could explain coexistence of sensitive and resistant strains of commensal bacteria in the intestinal microbiota, after individuals are given antibiotics following infection.^36^ In this study, high fitness cost values were considered, assuming that resistant strains were 50% less transmissible than sensitive ones (c = 0.50, with parameter c defined in Figure 2A). Later, Davies et al. explicitly formalized the role of strain competition within the host, introducing multiple sub-compartments of resistant strain carriage and differential intra-host growth between resistant and sensitive strains.^26^ Including such within-host competition mechanisms successfully reproduced rates of aminopenicillin and fluoroquinolone resistance estimated in *E. coli* across a large range of antibiotic use rates for European countries, and allowed for more flexible co-existence than the commonly used multi-carriage model (“R-S-multi” model defined in Figure 2A).^26^ Here, they estimated lower resistance-associated transmission costs (c < 0.20 for both resistances in *E. coli*).^26^ One model also formalized plasmid-mediated acquisition of ESBL production in *E. coli*.^32^

Other modeling studies evaluated whether structure in the host population could impact and explain antibiotic resistance dynamics at the population level (e.g., by considering that different host groups were characterized by distinct antibiotic use rates and inter-groups connectivity). They explored different structures, including geographic regions,^29^ where spatial spillover led to weak observed antibiotic use-resistance associations in some regions, or host classes notably age.^35^ Overall, considering highly structured populations allowed for the coexistence of sensitive and resistant bacteria over variable antibiotic use rates in these two studies.^29,35^ Interestingly, another modeling study analyzing data from Ecuador showed that persistence of antibiotic resistant *E. coli* in communities inter-connected through travel could be explained either by high antibiotic consumption or high interconnectedness.^31^

#### Identifying and quantifying transmission routes

We identified six studies using models to characterize routes of transmission and/or quantify transmission parameters for antibiotic-resistant Enterobacterales. All these studies used the “R” model structure, considering only transmission of resistant bacteria (Table 1).

Knight et al. and Allel et al. explored the relative contribution of community versus healthcare settings in the overall transmission of antibiotic resistance.^38,39^ Their results both suggested that individuals from the general population mostly acquired resistant *E. coli* or other Gram-negative bacteria in the community, despite higher levels of antibiotic use and transmission in the hospital.^38,39^ Within the community, multiple transmission routes were investigated in Mughini-Gras et al., such as human, food, environment, farm and companion animals.^23^ Using a source-attribution model, they estimated that 60% of community carriage of ESBL-producing Enterobacterales originated from human-to-human transmission. Nevertheless, authors also estimated that, while they do not represent the main acquisition events, contribution from non-human sources is required to maintain observed levels of colonization in the community.^23^

In three studies, model fitting to individual antibiotic resistance carriage data provided parameter estimates for the transmission rate and carriage duration of ESBL-producing Enterobacterales.^25,33,37^ Results were highly dependent on the population considered in the models and are summarized in Table 2. Within households, probability of transmission from a colonized index case to other household members was estimated to be 12% from a returning traveler (95% confidence interval CI 5-18)^37^ and 67% from a discharged hospital patient (95% CI 38-88).^25^ In both studies, the estimated human-to-human transmission rate was in the order of 10^-3^ events per day, whereas the estimated background environmental acquisition rate was in the order of 10^-4^ events per day (Table 2). Interestingly, longer median carriage duration was estimated for discharged patients (267 days, 95% CI 173-433) and household members (111 days, 95% CI 56-217),^25^ than for returning travelers (30 days, 95% CI 29-33), possibly explaining differences in the estimated transmission probabilities.^37^ Within the general population, Van Den Bunt et al. estimated the human-to-human transmission rate to be 9×10^-4^ events per colonized individual per day (95% CI 6×10^-4^ - 10×10^-4^).^33^ In agreement with estimates from Haverkate et al,^25^ the median duration of carriage was estimated to be 128 days (95% CI 106-157).

**Table 2.** Transmission model parameter estimates for ESBL-producing Enterobacterales, obtained from individual-based models informed by longitudinal antibiotic resistance carriage data.

| **Study** | **Host population** | **Human-to-human transmission rate**<br>(95% confidence interval) | **Background environmental acquisition rate**<br>(95% confidence interval) | **Median carriage duration**<br>(95% confidence interval) |
| --- | --- | --- | --- | --- |
| **Arcilla et al. 2017** <sup>37</sup> | Returning travelers, within households | $1.3 \times 10^{-3}$ /colonized individual/day<br>($0.5 \times 10^{-3}$ - $2.4 \times 10^{-3}$) | $7.3 \times 10^{-4}$ /day<br>($5.4 \times 10^{-4}$ - $9 \times 10^{-4}$) | 30 days<br>(29-33) for returning travelers |
| **Haverkate et al. 2017 (x2)** <sup>25</sup> | Discharged patients, within household | $5.3 \times 10^{-3}$ /colonized individual/day<br>($2.5 \times 10^{-3}$ - $11 \times 10^{-3}$) | $1.5 \times 10^{-4}$ /day<br>($0.2 \times 10^{-4}$ - $3.9 \times 10^{-4}$) | 267 days<br>(173 - 433) for discharged patients<br>111 days<br>(56 - 217) for household members |
| **Van den Bunt et al. 2020** <sup>33</sup> | General population | $9 \times 10^{-4}$ /colonized individual/day<br>($6 \times 10^{-4}$ - $10 \times 10^{-4}$) | - | 128 days<br>(106 - 157) for healthy individuals |

#### Evaluating the impact of population-level interventions on antibiotic resistance dynamics in the community

Six studies evaluated the effectiveness of public health interventions at reducing antibiotic resistance rates in Enterobacterales in the community (Table 1). Interventions included: antibiotic use reduction in the hospital, the community, or both;^22,24,27,28,30^ antibiotic molecule replacement in the community;^34^ and person-to-person transmission reduction in the hospital,^24,27^ or in the community through hand hygiene.^28,30^

Among those, two modeling studies tested the theoretical impact of community-focused interventions. At both the household and population levels, modeling results suggested that hand hygiene was more effective than antibiotic use restriction to reduce ESBL-producing *E. coli* transmission.^28,30^ In another study, using data from a 2-year intervention in a county in Sweden, Sundqvist et al. analyzed the impact of the substitution of trimethoprim with other antibiotics on resistance rates in *E. coli* urinary isolates.^34^ They found a low reversibility of resistance, both from time-series analyses and a population-level mathematical model. They also estimated a relatively low fitness cost associated with trimethoprim resistance (c = 0.01-0.02, with parameter c defined in Figure 2A), likely contributing to this low reversibility.

Three models combining nosocomial and community transmission were used to compare the impact of interventions across these two settings.^22,24,27^ MacFadden et al. fitted a transmission model to observed proportions of ESBL production among *E. coli* over time in Sweden.^22^ They predicted that a 20% decrease in antibiotic use in the community led to greater reductions in ESBL prevalence in both settings (community and hospital) than a same 20% decrease in hospital antibiotic use. Concerning *K pneumoniae*, two studies fitted their model to surveillance data of ESBL and carbapenem resistance rates over time in several European countries. In contrast to above estimates for *E. coli*, results from Kachalov et al. suggested that reducing nosocomial transmission and antibiotic consumption in hospitals had the strongest impact on *K. pneumoniae* resistance prevalence overall.^24^ Salazar-Vizcaya et al. predicted that a 50% decrease in consumption of all antibiotic classes in both hospitals and the community would lead to a small but significant decline in the prevalence of resistant *K. pneumoniae* in upcoming years in both settings, whereas a 50% decrease in the in-hospital transmission rate would lead to a decline only in hospital.^27^

### Empirical data for statistical inference of parameters

Of the 18 reviewed studies, half inferred model parameters by fitting model simulations to antibiotic resistance data collected in human populations (Table 3). Among those, data either described individual-level carriage or aggregated region- or country-level cases of infection.

**Table 3.**
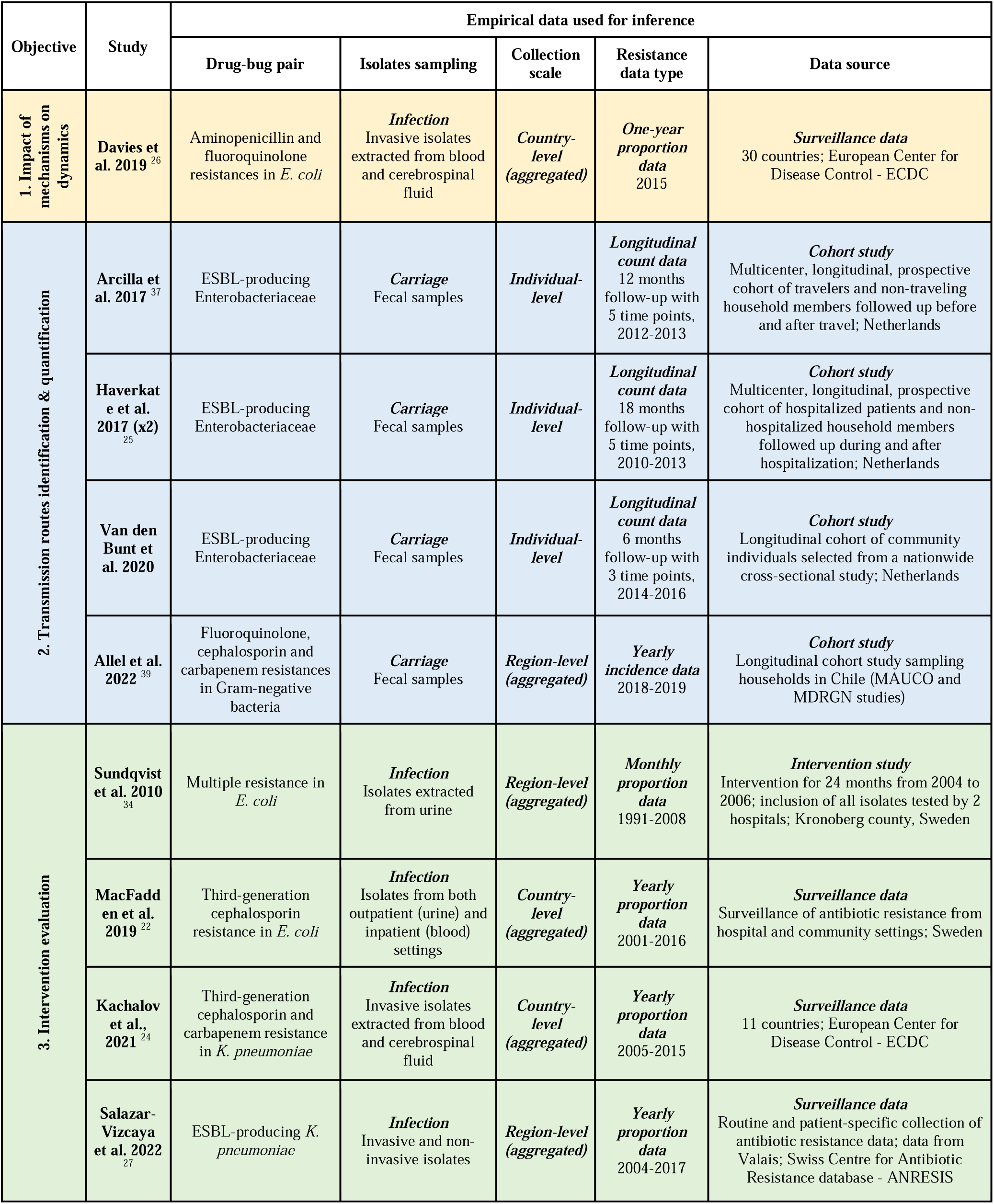
Details on empirical data used for inference in the reviewed modeling studies.

Antibiotic resistance carriage data were obtained from microbial analysis of fecal samples taken from individuals. Depending on the study, stool was sampled from healthy volunteers or from high-risk groups at regular intervals (days or months) to detect intestinal carriage of ESBL-producing Enterobacterales. The analysis of consecutive individual samples provides temporal information on the presence or absence of resistant bacteria, which can then be used to track bacterial acquisition or clearance. In three studies, longitudinal individual-level carriage data were accessed from cohort studies.^25,33,37^ These models were formalized using the “R” structure, where the acquisition and transmission of only resistant bacteria is described (Table 1). This is probably due to a lack of information on non-resistant bacteria carriage in the available data.

The other main type of data considered was aggregated counts of infections sensitive or resistant to a particular antibiotic, by unit of time at the region or country level. These data predominantly came from national surveillance systems, such as in Sweden or Switzerland,^22,27^ or at the European level.^24,26^ One study used aggregated infection cases from an intervention study.^34^ In these studies, the “R-S” or “R-S-multi” structures were usually chosen, considering the circulation of both resistant and non-resistant bacteria in the population. However, most of them considered only an infection stage, i.e. not considering carriage (Table 1).

The nine remaining studies did not estimate transmission parameters from data, relying instead on existing estimates from studies published in the literature. In one study, assumed parameters values were specific to Enterobacterales,^38^ while, for two others, values estimated for *S. pneumoniae* were considered.^29,35^ While these latter models were explicitly proposed to describe antibiotic resistance in generic commensal bacteria (as labeled “G” in Table 1), assumed resistance-associated fitness cost values were derived from *S. pneumoniae* and fixed to c = 0.168,^29^ or c = 0.04.^35^

## Discussion

In this systematic review, we identified and analyzed 18 mathematical modeling studies published between 1992 and 2023 that describe the transmission of antibiotic-resistant Enterobacterales in the community. Overall, models varied substantially in their structure, assumptions and focus, but nonetheless provided consistent insights by (i) confirming the significant role of community transmission in the overall burden of antibiotic-resistant Enterobacterales, (ii) investigating how various candidate mechanisms explained observed epidemiological trends, (iii) providing species-specific estimates of key parameters related to unobserved processes such as transmission and clearance and (iv) projecting how the effectiveness of public health interventions depended on settings, bacterial species, and types of resistance. Despite important contributions in the last decade, we identified a lack of modeling studies specifically designed for antibiotic-resistant Enterobacterales in the community, likely linked to a paucity of standardized data in this setting.

While the spread of antibiotic resistance is frequently associated with hospitals, which are environments with high rates of antibiotic use and reservoirs of multidrug-resistant organisms, results from the reviewed modeling studies highlighted the significant role of the community in driving the overall spread of antibiotic-resistant Enterobacterales.^22,38,39^ Epidemiological data also supported this hypothesis, showing that asymptomatic carriage of antibiotic-resistant Enterobacterales resulted mostly from frequent community-based acquisition events, at least for *E. coli*.^43,44^ Consequently, the community and hospitals likely both play significant but distinct roles in the global emergence and dissemination of resistance in Enterobacterales. Modeling their transmission within and between these two settings is thus highly relevant, and several of the reviewed studies did so.^22,24,27,28,32,38,39^ However, only two studies used detailed antibiotic resistant infection data specific to the two environments.^22,27^ In the future, extensive antibiotic resistance data collection in the community, through general practitioners or outpatient microbiological laboratories, should enable setting-specific model parametrization and therefore help to better characterize the contribution of each setting to transmission.

Robust modelling of antibiotic-resistant Enterobacterales transmission in the community requires accounting for heterogeneity in risks of transmission or acquisition, and carriage duration. Modeling studies have quantified the high risks of transmission of antibiotic-resistant Enterobacterales from specific groups such as discharged hospital patients and colonized infants to healthy members of their households.^25,30^ From epidemiological studies, it has been shown that travel to high-prevalence areas, including certain countries in Africa and Asia, was an important risk factor for resistant Enterobacterales acquisition.^20,46^ However, despite this high acquisition risk, both model and epidemiological studies suggested that returning travelers might be associated with a low subsequent transmission risk in households,^37^ possibly due to a short carriage duration.^37,47,48^ Future research should further investigate the traveler’s role in spreading resistant Enterobacterales locally: while the rapid global spread of high-risk multidrug-resistant clones is clearly linked to travel,^49^ it is unclear to what extent population movement contributes to community transmission dynamics once a clone has become established in a given region. Metapopulation models are powerful tools to account for impacts of such demographic and spatial heterogeneity on transmission, and could be harnessed to assess the net impact of public health interventions targeting specific sub-regions or sub-populations to reduce overall antibiotic resistance levels.^50^ However, the development of such models incorporating multiple sub-populations and risk profiles requires access to fine-grained epidemiological data, including data on asymptomatic carriage, infection and antibiotic use stratified by age, location, and other relevant covariates.

Several of the reviewed studies were not species-specific but rather considered generic Enterobacterales or intestinal commensals,^29,35,36,39^ with two of them using epidemiological parameters previously estimated for *S. pneumoniae*.^29,35^ However, Enterobacterales and *S. pneumoniae* do not share the same ecological niche within the host (mainly gut vs upper respiratory tract) and differ in primary transmission routes (direct or indirect orofecal vs airborne transmission), antibiotic resistance mechanisms (e.g., efflux pumps removing antibiotics from the cell in some strains of macrolide-resistant *S. pneumoniae* versus the production of antibiotic-degrading enzymes among ESBL-producing Enterobacterales) or in the fraction of the human population that are carriers.^51,52^ Such specificities suggest that commonly used pneumococcus models may have limited relevance for the study of Enterobacterales.^53^ Importantly, even within Enterobacterales, strong heterogeneity exists between species: while most individuals carry *E. coli, K. pneumoniae* is carried in less than 50% of the general healthy population,^54^ and non-*E. coli* Enterobacterales have been associated with greater transmissibility than *E. coli*.^55,56^ In addition, most pneumococcus models estimated strong serotype-dependent fitness costs associated with resistance, resulting in reduced transmission capacity of resistant strains compared with sensitive ones.^57,58^ While there is increasing evidence of resistance-associated fitness costs in Enterobacterales *in vitro*,^59,60^ such estimates at the epidemiological level are rare. Such costs may be expected to vary not only across *E. coli, K. pneumoniae* and other Enterobacterales, but also depending on whether a specific resistance mechanism is mutation- or horizontally-acquired.^61,62^ Improving our knowledge on such key parameters across bacterial species and resistance mechanisms, from *in vivo* and epidemiological studies, will be needed to develop more biologically relevant models. This will help to more consistently reproduce antibiotic resistance dynamics and better assess the expected impact of public health interventions on antibiotic resistance.^34^

As commensals, Enterobacterales thrive in the complex ecological environment that is the human gut. In this environment, myriad ecological phenomena occur, such as bacterial interactions, niche competition and HGT. Such phenomena have been suggested to influence the acquisition, carriage, and pathogenesis of antibiotic-resistant Enterobacterales.^63^ For example, dysbiosis following antibiotic use has been shown to reduce colonization resistance, the degree of protection conferred by the microbiome against the invasion of new and potentially antibiotic-resistant strains, or the overgrowth of pre-existing resistant strains.^64^ Regardless of potential impacts of antibiotics, host heterogeneity in microbiota composition likely also modulates carriage risk and duration.^65^ Further, HGT is known to contribute to the endogenous acquisition of resistance genes among resident bacteria, in part driving the observed prevalence of antibiotic-resistant Enterobacterales carriage.^62^ Many of the models included in this review have used structures that implicitly assume some degree of within-host ecological interaction between Enterobacterales and other microbiota. However, explicit data-driven quantification of such interactions has been scarce, and a previous review has highlighted that impacts of microbiota and HGT have been, until now, only marginally included in population-level models of Enterobacterales epidemiology. ^66^

To maintain sustained antibiotic-resistant Enterobacterales transmission in the community, reviewed models supported the hypothesis that non-human-source transmission events are relevant, including contact with animals or contaminated water, especially in low-income contexts.^23^ ^31,39^ However, approaches used to infer these transmission routes depended heavily on input data.^67^ The appropriate design of community cohorts where humans, animals and the environment could be sampled in a controlled manner, collecting both genomic and epidemiological data, would help better infer the relative input of potential environmental transmission routes. Methodological developments will also be essential to better incorporate such genomic data within epidemiological transmission models.

This review also illustrated the importance of adapting model structure to the scientific question being asked and its specific context. Models aimed at quantifying transmission routes all used the “R” structure, while those aimed at assessing the impact of different mechanisms on antibiotic resistance dynamics frequently used the “R-S” and “R-S-multi” structures (Table 1). Such structural choices may have important consequences for model predictions. On one hand, the “R-S” structure, which assumes a resistance-associated fitness cost, predicts a key impact of antibiotic use on the invasion, maintenance, and transmission of competing sensitive and resistant strains in populations. However, accurate modelling of closely related strains requires knowledge about the degree to which these strains compete and co-colonize the same ecological niche. On the other hand, “R” structure models do not account for such ecological competition and thus fail to capture one of the driving forces of Enterobacterales epidemiology.^26^ However, this simplified model structure may nonetheless be appropriate in certain contexts, for example when data on sensitive bacteria are unavailable and antibiotic selection for resistance can be incorporated parametrically.

Importantly, estimating key parameters to characterize antibiotic-resistant Enterobacterales transmission requires access to and analysis of appropriate data. In this review, 9 out of the 18 studies fitted their model to empirical collected data from human populations. When data were used, isolate metadata were often not detailed, impeding the identification of the sample’s source (hospital or community) and type (carriage or infection). Yet such information is critical to correctly parametrize models. In many countries, hospital infection reporting is done routinely, providing extensive antibiotic susceptibility data on invasive isolates.^68^ In contrast, infection and carriage surveillance in the community is more difficult to put in place, as antibiotic susceptibility testing is not done routinely.^68^ This leads to scarce available data on Enterobacterales epidemiology in the community, especially for carriage. Future availability of robust carriage data in the community, through cohorts or cross-sectional studies, will be key to better inform models and further disentangle the role of asymptomatic colonization in resistance spread.

To conclude, our study provided a detailed summary of epidemiological insights provided by published modeling studies of antibiotic-resistant Enterobacterales transmission in the community setting. We highlighted that the available modeling literature is still scarce compared to the one focusing on other bacterial species or on hospital transmission.^13,69^ The continued integration of hypothesis driven mechanisms and relevant epidemiological data into community-based transmission models will be necessary to better formalize the acquisition, colonization, and transmission of these bacteria within the general population. Open challenges in the field are, among others, to include with more precision the within-host processes in population-level models, to better integrate genomic and epidemiological data and to explore One Health transmission routes (Figure 3). Moreover, we advocate for extensive collection of antibiotic resistance carriage data in the community and the standardization of clinical bacterial isolate metadata. Such considerations will be instrumental in improving our understanding of the ecology and epidemiology of antibiotic-resistant Enterobacterales. In turn, this will be key to support the design of efficient public health interventions (hygiene measures, antibiotic prescribing reductions, wastewater treatment, vaccines, etc.), and identifying optimal target populations (high-risk individuals, households, communities, etc.) to reduce antibiotic resistance.

**Figure 3.**
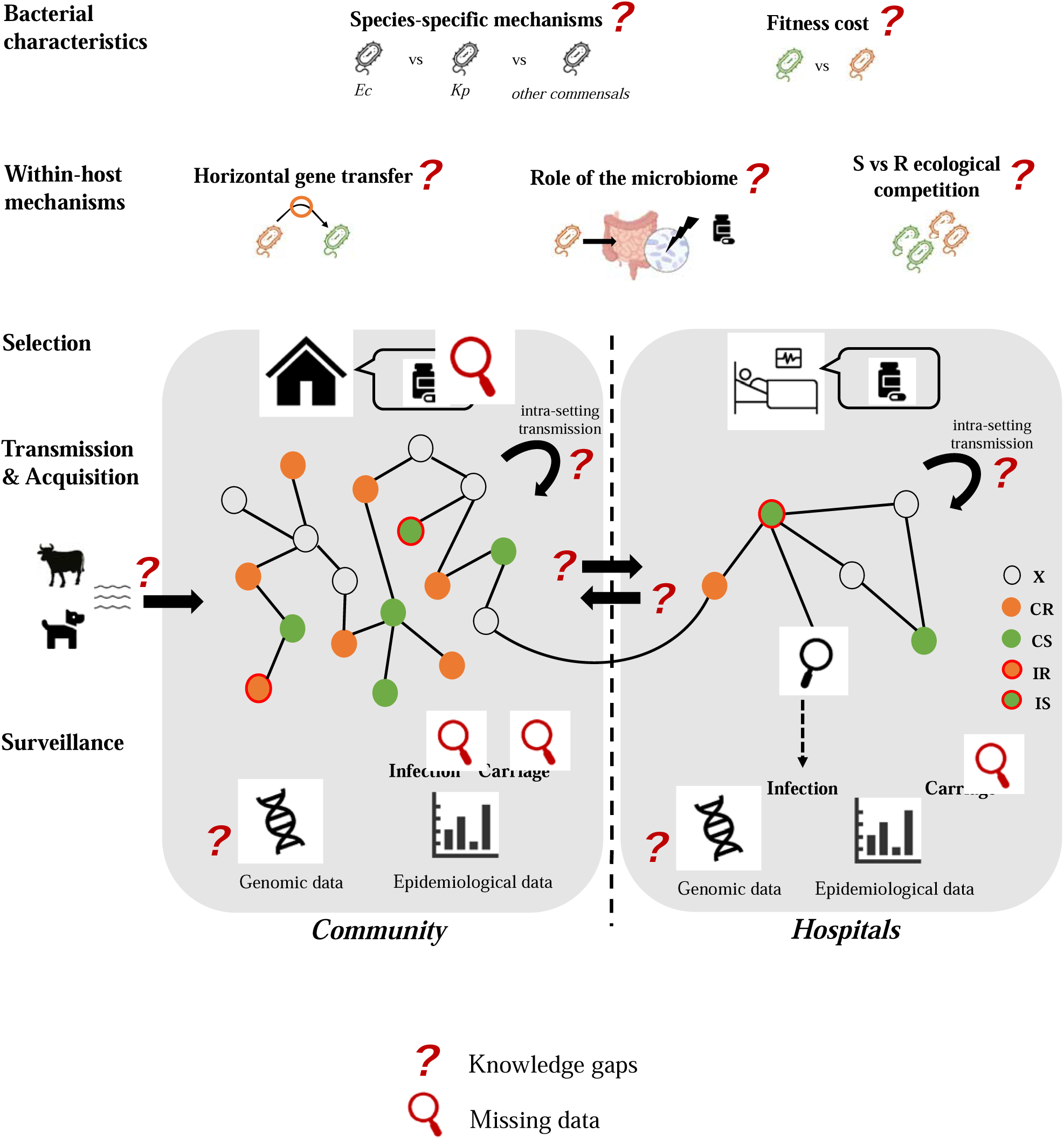
Modeling antibiotic-resistant Enterobacterales transmission between humans in a comprehensive framework: knowledge gaps and missing data. ***Bacterial characteristics.*** Biological and epidemiological characteristics of Enterobacterales differ from one another and from other important community-associated bacteria such as *S. pneumoniae*. *E. coli* and *K. pneumoniae*, for instance, are known to differ in terms of their transmissibility, carriage duration, and virulence properties. Different antibiotic-bacteria combinations also have specific resistance mechanisms and potential resistance-associated fitness costs. Model structure and parameterization should incorporate such biological knowledge and hypotheses. **Within-host mechanisms.** Most Enterobacterales are intestinal commensals, living in complex ecosystems. The role of the microbiota in colonization, infection and transmission should be further integrated into models, as well as the potential of horizontal gene transfer to accelerate the spread of resistance-conferring mobile genetic elements through bacterial and, in turn, human populations. **Selection.** Heterogeneous antibiotic use between populations (community vs hospitals; older vs younger individuals; different regions) might shape antibiotic resistance patterns in Enterobacterales, and more detailed data on specific consumption behaviors, especially in the community, are needed to better inform models. **Transmission & acquisition.** In addition to their roles as human intestinal symbionts, Enterobacterales circulate extensively among diverse vertebrate hosts and may persist for extended periods in environmental reservoirs. Models accounting for transmission from specific at-risk populations (e.g. discharged hospital patients) or non-human sources (e.g. pets, food) may help to elucidate transmission drivers in the community. **Surveillance.** High quality, standardized surveillance data from community settings are lacking, posing great challenges for accurate modeling. Specific transmission parameters should be informed by data specifically collected from resistant Enterobacterales, for both infection and carriage. Moreover, methodological development is needed to better incorporate genomic data into population-level epidemiological models. **Bacteria: Ec**: *Escherichia coli*; **Kp**: *Klebsiella pneumoniae*. **Compartments: X**: uncolonized individuals**; CS**: individuals colonized with sensitive strain**; CR**: individuals colonized with resistant strain**; IS**: individuals infected with sensitive strain**; IR**: individuals infected with resistant strain.

## Data Availability

All data produced in the present work are contained in the manuscript.

## Acknowledgments

We thank David RM Smith for critical feedback on the manuscript.

## Funding

ER’s work was funded by an independent research Pfizer Global Medical Grant (number 57504809). LO and PG’s work was funded by the French Government’s “Investissement d’Avenir” program, Laboratoire d’Excellence “Integrative Biology of Emerging Infectious Diseases” (Grant ANR-10-LABX-62-IBEID).

## Availability of data and materials

Not applicable.

## Authors’ contribution

ER, PG and LO designed the study. ER identified and analyzed the data. ER, PG and LO reviewed the data and wrote the manuscript. All authors read and approved the final manuscript.

## Declaration of interest

None.

## Notes

### Competing Interest Statement

The authors have declared no competing interest.

### Funding Statement

ER work was funded by an independent research Pfizer Global Medical Grant (number 57504809). LO and PG work was funded by the French Government Investissement d Avenir program Laboratoire d Excellence Integrative Biology of Emerging Infectious Diseases (Grant ANR-10-LABX-62-IBEID).

## Bibliography

1 W H O. Global action plan on antimicrobial resistance. 2015. http://www.emro.who.int/health-topics/drug-resistance/global-action-plan.html (accessed Sept 20, 2021).

2 Rahbe E, Watier L, Guillemot D, Glaser P, Opatowski L. Determinants of worldwide antibiotic resistance dynamics across drug-bacterium pairs: a multivariable spatial-temporal analysis using ATLAS. Lancet Planet Health 2023; 7: e547–57.

3 Antimicrobial Resistance Collaborators. Global burden of bacterial antimicrobial resistance in 2019: a systematic analysis. Lancet 2022; 399: 629–55.

4 Jenkins C, Rentenaar RJ, Landraud L, Brisse S. Enterobacteriaceae. In: Infectious Diseases. Elsevier, 2017: 1565–1578.e2.

5 Schwaber MJ, Carmeli Y. Carbapenem-resistant Enterobacteriaceae: a potential threat. JAMA 2008; 300: 2911–3.

6 Abban MK, Ayerakwa EA, Mosi L, Isawumi A. The burden of hospital acquired infections and antimicrobial resistance. Heliyon 2023; 9: e20561.

7 Bezabih YM, Sabiiti W, Alamneh E, et al. The global prevalence and trend of human intestinal carriage of ESBL-producing Escherichia coli in the community. J Antimicrob Chemother 2021; 76: 22–9.

8 Woerther P-L, Burdet C, Chachaty E, Andremont A. Trends in human fecal carriage of extended-spectrum β-lactamases in the community: toward the globalization of CTX-M. Clin Microbiol Rev 2013; 26: 744–58.

9 Bonten M, Johnson JR, van den Biggelaar AHJ, et al. Epidemiology of Escherichia coli Bacteremia: A Systematic Literature Review. Clin Infect Dis 2021; 72: 1211–9.

10 Pitout JDD, Nordmann P, Laupland KB, Poirel L. Emergence of Enterobacteriaceae producing extended-spectrum beta-lactamases (ESBLs) in the community. J Antimicrob Chemother 2005; 56: 52–9.

11 Opatowski L, Guillemot D, Boëlle P-Y, Temime L. Contribution of mathematical modeling to the fight against bacterial antibiotic resistance. Curr Opin Infect Dis 2011; 24: 279–87.

12 Bonten MJ, Austin DJ, Lipsitch M. Understanding the spread of antibiotic resistant pathogens in hospitals: mathematical models as tools for control. Clin Infect Dis 2001; 33: 1739–46.

13 Niewiadomska AM, Jayabalasingham B, Seidman JC, et al. Population-level mathematical modeling of antimicrobial resistance: a systematic review. BMC Med 2019; 17: 81.

14 Ramsay DE, Invik J, Checkley SL, Gow SP, Osgood ND, Waldner CL. Application of dynamic modelling techniques to the problem of antibacterial use and resistance: a scoping review. Epidemiol Infect 2018; 146: 2014–27.

15 Chatterjee A, Modarai M, Naylor NR, et al. Quantifying drivers of antibiotic resistance in humans: a systematic review. Lancet Infect Dis 2018; 18: e368–78.

16 Hilty M, Betsch BY, Bögli-Stuber K, et al. Transmission dynamics of extended-spectrum β-lactamase-producing Enterobacteriaceae in the tertiary care hospital and the household setting. Clin Infect Dis 2012; 55: 967–75.

17 Rodríguez-Baño J, López-Cerero L, Navarro MD, Díaz de Alba P, Pascual A. Faecal carriage of extended-spectrum beta-lactamase-producing Escherichia coli: prevalence, risk factors and molecular epidemiology. J Antimicrob Chemother 2008; 62: 1142–9.

18 Despotovic M, de Nies L, Busi SB, Wilmes P. Reservoirs of antimicrobial resistance in the context of One Health. Curr Opin Microbiol 2023; 73: 102291.

19 Ny S, Löfmark S, Börjesson S, et al. Community carriage of ESBL-producing Escherichia coli is associated with strains of low pathogenicity: a Swedish nationwide study. J Antimicrob Chemother 2017; 72: 582–8.

20 Raffelsberger N, Buczek DJ, Svendsen K, et al. Community carriage of ESBL-producing Escherichia coli and Klebsiella pneumoniae: a cross-sectional study of risk factors and comparative genomics of carriage and clinical isolates. mSphere 2023; : e0002523.

21 Godijk NG, Bootsma MCJ, Bonten MJM. Transmission routes of antibiotic resistant bacteria: a systematic review. BMC Infect Dis 2022; 22: 482.

22 MacFadden DR, Fisman DN, Hanage WP, Lipsitch M. The Relative Impact of Community and Hospital Antibiotic Use on the Selection of Extended-spectrum Beta-lactamase-producing Escherichia coli. Clin Infect Dis 2019; 69: 182–8.

23 Mughini-Gras L, Dorado-García A, van Duijkeren E, et al. Attributable sources of community-acquired carriage of Escherichia coli containing β-lactam antibiotic resistance genes: a population-based modelling study. Lancet Planet Health 2019; 3: e357–69.

24 Kachalov VN, Nguyen H, Balakrishna S, et al. Identifying the drivers of multidrug-resistant Klebsiella pneumoniae at a European level. PLoS Comput Biol 2021; 17: e1008446.

25 Haverkate MR, Platteel TN, Fluit AC, et al. Quantifying within-household transmission of extended-spectrum β-lactamase-producing bacteria. Clin Microbiol Infect 2017; 23: 46.e1–46.e7.

26 Davies NG, Flasche S, Jit M, Atkins KE. Within-host dynamics shape antibiotic resistance in commensal bacteria. Nat Ecol Evol 2019; 3: 440–9.

27 Salazar-Vizcaya L, Atkinson A, Kronenberg A, et al. The impact of public health interventions on the future prevalence of ESBL-producing Klebsiella pneumoniae: a population based mathematical modelling study. BMC Infect Dis 2022; 22: 487.

28 Talaminos A, López-Cerero L, Calvillo J, Pascual A, Roa LM, Rodríguez-Baño J. Modelling the epidemiology of Escherichia coli ST131 and the impact of interventions on the community and healthcare centres. Epidemiol Infect 2016; 144: 1974–82.

29 Olesen SW, Lipsitch M, Grad YH. The role of “spillover” in antibiotic resistance. Proc Natl Acad Sci USA 2020; 117: 29063–8.

30 Kardaś-Słoma L, Yazdanpanah Y, Perozziello A, et al. Hand hygiene improvement or antibiotic restriction to control the household transmission of extended-spectrum β-lactamase-producing Escherichia coli: a mathematical modelling study. Antimicrob Resist Infect Control 2020; 9: 139.

31 Eisenberg JNS, Goldstick J, Cevallos W, et al. In-roads to the spread of antibiotic resistance: regional patterns of microbial transmission in northern coastal Ecuador. J R Soc Interface 2012; 9: 1029–39.

32 Godijk NG, Bootsma MCJ, van Werkhoven HC, et al. Does plasmid-based beta-lactam resistance increase E. coli infections: Modelling addition and replacement mechanisms. PLoS Comput Biol 2022; 18: e1009875.

33 van den Bunt G, Fluit AC, Bootsma MCJ, et al. Dynamics of Intestinal Carriage of Extended-Spectrum Beta-lactamase-Producing Enterobacteriaceae in the Dutch General Population, 2014-2016. Clin Infect Dis 2020; 71: 1847–55.

34 Sundqvist M, Geli P, Andersson DI, et al. Little evidence for reversibility of trimethoprim resistance after a drastic reduction in trimethoprim use. J Antimicrob Chemother 2010; 65: 350–60.

35 Blanquart F, Lehtinen S, Lipsitch M, Fraser C. The evolution of antibiotic resistance in a structured host population. J R Soc Interface 2018; 15. DOI:10.1098/rsif.2018.0040.

36 Austin DJ, Kakehashi M, Anderson RM. The transmission dynamics of antibiotic-resistant bacteria: the relationship between resistance in commensal organisms and antibiotic consumption. Proc Biol Sci 1997; 264: 1629–38.

37 Arcilla MS, van Hattem JM, Haverkate MR, et al. Import and spread of extended-spectrum β-lactamase-producing Enterobacteriaceae by international travellers (COMBAT study): a prospective, multicentre cohort study. Lancet Infect Dis 2017; 17: 78–85.

38 Knight GM, Costelloe C, Deeny SR, et al. Quantifying where human acquisition of antibiotic resistance occurs: a mathematical modelling study. BMC Med 2018; 16: 137.

39 Allel K, Goscé L, Araos R, et al. Transmission of gram-negative antibiotic-resistant bacteria following differing exposure to antibiotic-resistance reservoirs in a rural community: a modelling study for bloodstream infections. Sci Rep 2022; 12: 13488.

40 Spicknall IH, Foxman B, Marrs CF, Eisenberg JNS. A modeling framework for the evolution and spread of antibiotic resistance: literature review and model categorization. Am J Epidemiol 2013; 178: 508–20.

41 Johnson JR, Owens K, Gajewski A, Clabots C. Escherichia coli colonization patterns among human household members and pets, with attention to acute urinary tract infection. J Infect Dis 2008; 197: 218–24.

42 Leimbach A, Hacker J, Dobrindt U. E. coli as an all-rounder: the thin line between commensalism and pathogenicity. Curr Top Microbiol Immunol 2013; 358: 3–32.

43 Herindrainy P, Rabenandrasana MAN, Andrianirina ZZ, et al. Acquisition of extended spectrum beta-lactamase-producing enterobacteriaceae in neonates: A community based cohort in Madagascar. PLoS One 2018; 13: e0193325.

44 Madigan T, Johnson JR, Clabots C, et al. Extensive Household Outbreak of Urinary Tract Infection and Intestinal Colonization due to Extended-Spectrum β-Lactamase-Producing Escherichia coli Sequence Type 131. Clin Infect Dis 2015; 61: e5–12.

45 García-Rey C, Fenoll A, Aguilar L, Casal J. Effect of social and climatological factors on antimicrobial use and Streptococcus pneumoniae resistance in different provinces in Spain. J Antimicrob Chemother 2004; 54: 465–71.

46 Otter JA, Natale A, Batra R, et al. Individual- and community-level risk factors for ESBL Enterobacteriaceae colonization identified by universal admission screening in London. Clin Microbiol Infect 2019; 25: 1259–65.

47 Ling W, Peri AM, Furuya-Kanamori L, Harris PNA, Paterson DL. Carriage Duration and Household Transmission of Enterobacterales Producing Extended-Spectrum Beta-Lactamase in the Community: A Systematic Review and Meta-Analysis. Microb Drug Resist 2022; published online June 21. DOI:10.1089/mdr.2022.0035.

48 Ruppé E, Armand-Lefèvre L, Estellat C, et al. High Rate of Acquisition but Short Duration of Carriage of Multidrug-Resistant Enterobacteriaceae After Travel to the Tropics. Clin Infect Dis 2015; 61: 593–600.

49 van der Bij AK, Pitout JDD. The role of international travel in the worldwide spread of multiresistant Enterobacteriaceae. J Antimicrob Chemother 2012; 67: 2090–100.

50 Krieger MS, Denison CE, Anderson TL, Nowak MA, Hill AL. Population structure across scales facilitates coexistence and spatial heterogeneity of antibiotic-resistant infections. PLoS Comput Biol 2020; 16: e1008010.

51 Bogaert D, van Belkum A, Sluijter M, et al. Colonisation by Streptococcus pneumoniae and Staphylococcus aureus in healthy children. Lancet 2004; 363: 1871–2.

52 Tenaillon O, Skurnik D, Picard B, Denamur E. The population genetics of commensal Escherichia coli. Nat Rev Microbiol 2010; 8: 207–17.

53 Lipsitch M, Colijn C, Cohen T, Hanage WP, Fraser C. No coexistence for free: neutral null models for multistrain pathogens. Epidemics 2009; 1: 2–13.

54 Chang D, Sharma L, Dela Cruz CS, Zhang D. Clinical Epidemiology, Risk Factors, and Control Strategies of Klebsiella pneumoniae Infection. Front Microbiol 2021; 12: 750662.

55 Duval A, Obadia T, Boëlle P-Y, et al. Close proximity interactions support transmission of ESBL-K. pneumoniae but not ESBL-E. coli in healthcare settings. PLoS Comput Biol 2019; 15: e1006496.

56 Denkel LA, Maechler F, Schwab F, et al. Infections caused by extended-spectrum β-lactamase-producing Enterobacterales after rectal colonization with ESBL-producing Escherichia coli or Klebsiella pneumoniae. Clin Microbiol Infect 2020; 26: 1046–51.

57 Maher MC, Alemayehu W, Lakew T, et al. The fitness cost of antibiotic resistance in Streptococcus pneumoniae: insight from the field. PLoS One 2012; 7: e29407.

58 de Cellès MD, Pons-Salort M, Varon E, et al. Interaction of vaccination and reduction of antibiotic use drives unexpected increase of pneumococcal meningitis. Sci Rep 2015; 5: 11293.

59 Melnyk AH, Wong A, Kassen R. The fitness costs of antibiotic resistance mutations. Evol Appl 2015; 8: 273–83.

60 Vanacker M, Lenuzza N, Rasigade J-P. The fitness cost of horizontally transferred and mutational antimicrobial resistance in Escherichia coli. Front Microbiol 2023; 14: 1186920.

61 Enne VI, Delsol AA, Davis GR, Hayward SL, Roe JM, Bennett PM. Assessment of the fitness impacts on Escherichia coli of acquisition of antibiotic resistance genes encoded by different types of genetic element. J Antimicrob Chemother 2005; 56: 544–51.

62 MacLean RC, San Millan A. The evolution of antibiotic resistance. Science 2019; 365: 1082– 3.

63 Lamberte LE, van Schaik W. Antibiotic resistance in the commensal human gut microbiota. Curr Opin Microbiol 2022; 68: 102150.

64 Pamer EG. Resurrecting the intestinal microbiota to combat antibiotic-resistant pathogens. Science 2016; 352: 535–8.

65 Chang Q, Lipsitch M, Hanage WP. Impact of Host Heterogeneity on the Efficacy of Interventions to Reduce Staphylococcus aureus Carriage. Infect Control Hosp Epidemiol 2016; 37: 197–204.

66 Leclerc QJ, Lindsay JA, Knight GM. Mathematical modelling to study the horizontal transfer of antimicrobial resistance genes in bacteria: current state of the field and recommendations. J R Soc Interface 2019; 16: 20190260.

67 Mughini-Gras L, Kooh P, Augustin J-C, et al. Source attribution of foodborne diseases: potentialities, hurdles, and future expectations. Front Microbiol 2018; 9: 1983.

68 Johnson AP. Surveillance of antibiotic resistance. *Philos Trans R Soc Lond B*, Biol Sci 2015; 370: 20140080.

69 Assab R, Nekkab N, Crépey P, et al. Mathematical models of infection transmission in healthcare settings: recent advances from the use of network structured data. Curr Opin Infect Dis 2017; 30: 410–8.

